# The global pattern of centenarians highlights deep problems in demography

**DOI:** 10.1101/2024.09.06.24313170

**Authors:** Saul Justin Newman

## Abstract

Accurate age data is fundamental to medicine, social sciences, epidemiology, and good government. However, recent and heavily disputed debates on data quality have raised questions on the accuracy of demographic data at older ages. Here, we catalogue late-life survival patterns of every country in the world from 1970-2021 using comprehensive estimates of old-age populations provided by global governments and curated by the United Nations. Analysis of 236 nations or states across 51 years reveals that late-life survival data is dominated by anomalies at all scales and in all time periods. Life expectancy at age 100 and late-life survival from ages 80 to 100+, which we term centenarian attainment rate, is highest in a seemingly random assortment of states. The top 10 ‘blue zone’ regions with the best survival to ages 100+ routinely includes Thailand, Kenya and Malawi – respectively now 212^th^ and 202^nd^ in the world for life expectancy, the non-self-governing territory of Western Sahara, and Puerto Rico where birth certificates are so unreliable they were recently declared invalid as a legal document. These anomalous rankings are conserved across long time periods and multiple non-overlapping cohorts, and do not seem to be sampling effects. Instead these patterns suggest a persistent inability, even for nation-states or global organisations, to detect or measure error rates in human age data, with troubling implications for epidemiology, demography, and medicine.

## Introduction

Chronological age is a fundamental metric in medicine and public health, constituting one of the single most important and informative risk factors for mortality and morbidity across virtually every population and disease. Age-specific mortality rates and their derivative measures, calculated directly from aggregated age data, likewise form a core international metric of human health^1–4^. These metrics are used by global governments to project and plan for future old-age survival, affecting the long-term planning for infrastructure and spending in healthcare^5–7^, the hedging of longevity risk^8^, and the setting of pension rates that allocate trillions of dollars to provide for future old-age populations^9^.

However, the measurement of human ages, and by extension age-specific rates of any quantity, relies almost universally upon a single measurement system: the globally-incomplete^10,11^ paperwork-based system of documentary evidence known as vital registration^10–13^. Despite age being the single most important risk factor for human health, along with gender, there has been no accurate and independently metric to validate human age measurements. If a developmentally mature person walks into a clinical setting with no paperwork, for example, there has been no independent or reproducible test available to measure their chronological age^14^. As such, if age-based paperwork consistently records an incorrect age, there is no method by which that error can be detected^14^ because there is, or rather has been^15–18^, no independently reproducible scientific method available for discovering such errors.

As a result, globally diverse document-based systems of vital registration are not subject to any document-independent technical validation or calibration. Systematic errors or error-generating processes that modify age records, from heavily biased or systemic errors to simple typographic mistakes, can therefore remain undetected indefinitely.

Despite some scepticism on the reliability of age data^19–21^ this situation has been long ignored: first on the basis of an untestable assumption that such errors must be rare^22^, and second on the seemingly reasonable statistical grounds that – if vital registration errors are assumed to be sufficiently rare and random – they may be safely ignored by fitting random error terms within a statistical model.

Recent theoretical work has shown that neither case seems to be a valid assumption^23^ especially at older ages^19,23^. In survival processes, age-coding errors accumulate non-randomly with age — even when initial rates of error are vanishingly low, symmetrically distributed, and random — through a process that can substantially distort late-life data^23,24^ and massively inflate the frequency of errors at certain ages (see accompanying theoretical paper).

The underlying theoretical reason is simple. Consider, for example, a population of one million fifty-year-old people, into which a hundred 40-year-olds are accidentally included through age-coding errors: an initial error rate of 0.01% or one in every ten thousand. The paperwork of these 40-year-olds accidentally records them as aged 50 years — a surprisingly common mistake^25,26^ – and these ‘young liar’ errors appear, officially and on paper, as 50-year-olds. As the two cohorts age, the ‘young liar’ errors are less than half as likely to die as the actual 50-year-olds — because they are biologically 10 years younger — and errors therefore constitute a growing fraction of the population with age. In typical human populations, error rates will grow at an approximately exponential rate with age due to the better survival of ‘young liars.’ By age 85 more than half of the population becomes errors, by age 100 ‘young liar’ errors constitute the entire population: a kind of error explosion caused by the asymmetrically better survival of ‘young liars.’

That is, because errors survive at higher rates than accurate data in survival processes, randomly distributed errors accumulate nonrandomly with age until they constitute the entire population. This process can cause initially rare random errors, of below 1 in 10,000 people, to accumulate in old-age data at extremely high frequencies within highly non-random distributions^23^.

Combined with the historical lack of paperwork-independent methods to validate and correct paper records, this simple theoretical process raises an uncomfortable possibility: that extreme age records may be dominated by undetected errors^23^. Here, we explore this possibility using the largest comprehensive survey of extreme-old populations in the world, assembled by the collective governments of United Nations (UN) member states, and curated by the United Nations Population Division into the World Population Prospects report^27^.

## Results

Data on the global population of centenarians was downloaded from 1950-2021 from the UN World Population Prospects 2022 edition^27^ for every UN member state and country in the world, and raw data used to calculate cohort survival rates from ages 80-84 to ages 100+ using the fmsb package^28^ in R version 4.3.3 (2024-02-29)^29^. These data reveal troubling patterns.

Even unmodified pre-calculation estimates, published directly by the United Nations and not subject to modification, display anomalies. For example, incongruous patterns are evident across the unadjusted UN estimates of life expectancy at age 100 – the number of additional years an average 100-year-old can expect to survive – for the most recently available data from 2021 (Fig S1; Table S1).

According to uncorrected UN metrics, the ‘blue zone’ regions with the highest late-life survival are heavily enriched for states with absent or unreliable birth certificates, states with no centralised government, communist dictatorships, and countries actively engaged in war or genocide. New Caledonia is the best country in the world for life expectancy at age 100, despite being ranked 51^st^ in the world for life expectancy at birth. New Caledonia was closely followed in the rankings by Puerto Rico and Thailand (Fig S1; Table S1). Puerto Rico was an especially unusual outcome for second place, as it suffers chronic problems with birth certificates being stolen, forged, sold online, or simply mis-filed or filled out with incorrect birthdates^30,31^. This situation is so severe that every birth certificate issued before 2010 was invalidated, cancelling their status as a legal document^30^, and the entire birth certification system restarted^30^. Inclusion of Puerto Rico as a leading state for survival past age 100, therefore, suggests that the data-cleaning efforts of leading demographers remains somewhat unable to adjust for obvious error-generating processes.

For 2021, the remaining states with high life expectancy at age 100+ includes a large number of colonial and post-colonial holdings with high rates of poverty relative to the national average, marking them out as regions of generally poor record-keeping and underfunded health systems, and simultaneously, desirable as locations to retire for rich internal migrants (Table S1). Both factors may inflate estimates of late-life survival, the first through uncorrected error processes and the second through old-age migration flows, which remain uncorrected in the UN statistics^32^.

These consistently anomalous results were reinforced when estimating more robust, long-term rates of cohort survival (Supplementary Code), To supplement UN estimates – that are largely model-based, cross-sectional, and apply to quinquennial age groups or open-ended age categories^27,33^ – we captured cohort survival rates from age 80-100+ using raw estimates of centenarian numbers available for every UN member state from 1950-2021.

Calculation of longitudinal cohort survival rates from age 80 to age 100+, which we term the centenarian attainment rate, reveals highly anomalous results across. Across 51 years of available data for 236 different states, the leading countries for centenarian attainment were over-represented for communist and post-communist states, states without birth certificates, dictatorships, and states at war (Table 1).

**Table 1.** Percent of 80-84-year-olds reaching age 100+ by country and year, shown for top 10 countries 1970-2021.

| Percent - Country |  |  |  |
| --- | --- | --- | --- |
| Rank | 1970 | 1980 | 1990 |
| 1 | 4.5 - Puerto Rico | 15.0 - Western Sahara | 9.8 - Malawi |
| 2 | 2.5 - Côte d'Ivoire | 4.7 - Puerto Rico | 7.2 - Mexico |
| 3 | 1.9 - Thailand | 3.6 - Thailand | 5.9 - Thailand |
| 4 | 1.7 - Bhutan | 3.1 - Paraguay | 3.0 - Puerto Rico |
| 5 | 1.7 - Paraguay | 2.8 - Jamaica | 3.0 - SVG |
| 6 | 1.7 - Bulgaria | 2.2 - SVG | 2.6 - Guatemala |
| 7 | 1.6 - SVG | 2.1 - Côte d'Ivoire | 2.0 - Montenegro |
| 8 | 1.3 - RF (USSR) | 2.0 - Malawi | 2.0 - Guadeloupe |
| 9 | 1.3 - Albania | 1.7 - Hong Kong SAR | 1.8 - Turkmenistan |
| 10 | 1.3 - Guyana | 1.6 - Guadeloupe | 1.7 - Hong Kong SAR |
| Rank | 2000 | 2010 | 2021 |
| 1 | 6.9 - Andorra | 5.9 - Malawi | 40.0 - Monaco |
| 2 | 6.4 - Malawi | 5.4 - Andorra | 8.2 - Thailand |
| 3 | 5.5 - Mexico | 5.2 - New Caledonia | 6.7 - Guadeloupe |
| 4 | 5.3 - Thailand | 5.1 - Western Sahara | 6.3 - Kenya |
| 5 | 5.3 - Hong Kong SAR | 4.9 - Thailand | 6.2 - Hong Kong SAR |
| 6 | 4.4 - Western Sahara | 4.9 - Monaco | 5.6 - Uruguay |
| 7 | 3.9 - Dominican Republic | 4.2 - Guadeloupe | 5.1 - Guam |
| 8 | 3.7 - Puerto Rico | 4.1 - El Salvador | 5.0 - Saint Martin (Fr) |
| 9 | 2.7 - Guadeloupe | 3.9 - Uruguay | 4.7 - Japan |
| 10 | 2.2 - Uruguay | 3.6 - Japan | 4.4 - Panama |

SVG = Saint Vincent and the Grenadines; RF (USSR) = Soviet Union

These irregularities persist into recent data (Fig 1). Arguably the most defensible ranking was Monaco, which was both the leading country for life expectancy and the leading country for centenarian attainment in 2021. The remainder of the rankings, however, appear to make somewhat less sense. The 2021 ‘blue zones’ – the countries with the highest centenarian attainment rates –include Thailand, Uruguay, and Kenya (as with life expectancies at age 100 above) alongside Monaco, Guadeloupe, and Hong Kong (Fig 1). In 2021, Kenya was simultaneously the 4^th^ best country in the world for centenarian attainment rates and the 213^th^ ranked region for life expectancy at birth (out of 236; Fig 1; Table 1). For context, mid-civil-war and ISIS-insurgency Syria was ranked 132^nd^ for average life expectancy in the same year.

**Figure 1.**
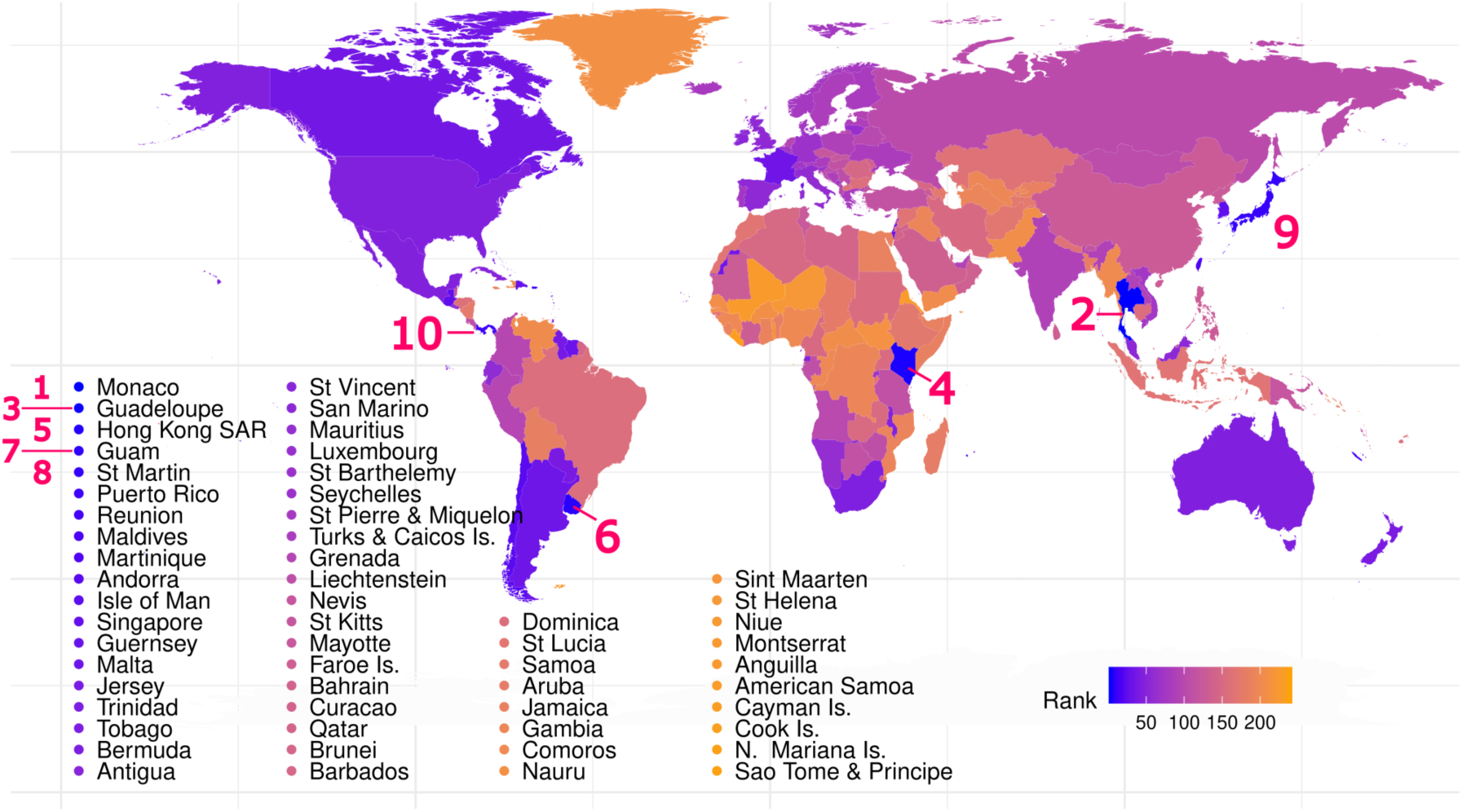
Centenarian attainment rates in 2021 United Nations data. Regions achieving the highest rate of cohort survival from ages 80 to 100+ years old – the centenarian attainment rate– are a highly irregular mix of states. For example, the top ten states globally for centenarian attainment (numbered) include both Monaco, with the highest life expectancy at birth in the world, and Kenya which is ranked 213^th^ in the world for life expectancy at birth. These rankings resemble unadjusted quinquennial UN figures for late-life survival – while being less model-dependent and less prone to sampling noise (Fig S1) – but have almost no resemblance to mortality or survival rates at any other age.

This dissonance continues across other UN member states, with Thailand, Guam, Panama, Uruguay respectively ranking 2^nd^, 7^th^, 10^th^, and 6^th^ for centenarian attainment, and 54^th^, 72^nd^, 79^th^, and 83^rd^ for average life expectancy (Table 1). Puerto Rico, regrettably, fell to 11^th^ place for centenarian attainment after its legislative clean-up of birth certificates^30^. The remaining countries with top 10 survival from ages 80-100+ were middle-to top-ranked countries for average life expectancy. This near-arbitrary set of countries does not seem to form a coherent grouping except, perhaps, as the collective outcome of an undetected error process.

The mismatch of late-life survival statistics with broad measurements of health and survivorship was not contained to top-ranked countries or specific years, and instead displayed consistently poor rank-concordance between life expectancy and centenarian attainment across all countries and years (Fig 2b). For example, Norway, Sweden, Lichtenstein, and Iceland – the latter has the most comprehensive long-term national records on earth – respectively had the 10^th^, 12^th^, 9^th^, and 15^th^ best life expectancies in the world in 2021. These countries also ranked, respectively, as 76^th^, 84^th^, 106^th^, and 79^th^ in the world for centenarian attainment.

**Figure 2.**
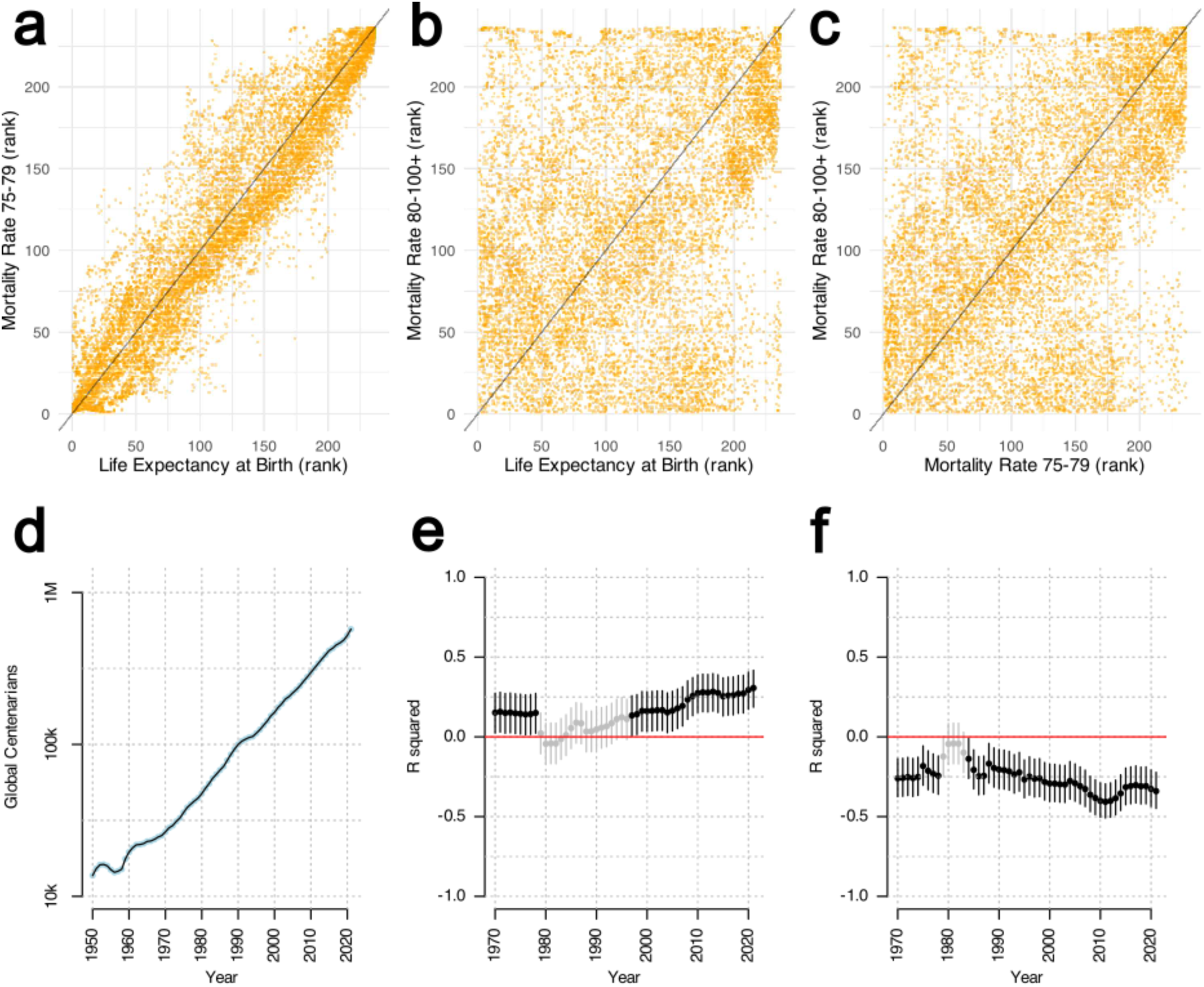
Persistently weak relationships between late-life and earlier-life survival rates. The relationship between mortality rates is highly rank-conserved across ages, even over substantial age gaps, as shown for example by (a) the high rank concordance between (cross-sectional) life expectancy at birth and mortality rates at ages 75-79. However, this relationship collapses in late life mortality data. Centenarian attainment rates — cohort mortality rates across ages 80-100 — are at best weakly correlated with life expectancy at birth (b), or with mortality rates at age 75-79 in the same cohort (c). Within-cohort concordance at younger ages is even worse. This randomness does not appear to reflect sample sizes. The estimated number of global centenarians has exploded at a log-linear rate since 1950 (d), alongside a huge global expansion in vital registration rates, yet this has not meaningfully improved the concordance between centenarian attainment and life expectancy (e), late-life mortality at age 75 (f), or mortality at any other age (Supplementary Code). Instead, concordance between early- and late-life survival remains within the standard error for data from 1970, after losing significance entirely during the early 1980s (grey points), and appears consistent with a hidden, uncorrected error-generating process.

Over time the most consistent global leaders in late-life survival, across all years, were Thailand – which never fell out of the top 10 rankings – and Puerto Rico, Malawi, and Hong Kong, all of which appeared more than 30 times in the top 10 rankings for centenarian attainment (Fig S2). The highest-ever records for centenarian attainment occurred in Monaco from 2014-2021 inclusive, with centenarian attainment rates so far above the other ranks that they appear to be error-driven outliers (Table 1). Monaco is followed closely by the non-self-governing territory of Western Sahara from 1979 until 1984 during a civil war, and then in Malawi from 1981-1992 during the peak of the AIDS pandemic. As of 2021 Malawi still ranks a respectable 46^th^ overall for centenarian attainment (Supplementary Code), yet this is a substantial decline in their rankings.

Remarkably, Malawi had been one of the top 3 states worldwide for centenarian attainment and late-life survival every year from 1983-2011 inclusive, despite struggling with a per capita GDP (PPP adjusted) below that of North Korea and a bottom-20 average life expectancy in the world throughout this entire period^27^.

The global ranking of UN member states in late-life survival did not correspond closely to metrics of survival at other ages (Fig 2; Fig S3). Normalising for period effects by using rankings within each year and calculating the rank-based correlation reveals concordance between different estimates of survival across all years in the UN data (Fig 2). Such rank-based comparisons remove period effects on overall mortality rates and reveal that life expectancy at age zero and mortality rates at age 70-74 (a seven-decade gap) are, as expected, closely correlated (Fig 2a; R^2^ = 0.93; p < 2e-16). Neither life expectancy at birth (Fig 2b; R^2^ = 0.33; p < 2e-16) or mortality rates at age 70-74 are, however, strongly correlated with survival at age 80-100+ (Fig 2c; Fig S3; R^2^ = 0.45; p < 2e-16; Supplementary Code) and routinely drop below the threshold for significance during the 1980s (Fig 2; Supplementary Code). In other words, late-life survival rankings do not predict rankings for mortality rates at age 70-74 or life expectancy rankings at age zero with any marked degree of accuracy (Fig 2) and appear to be largely uncoupled from estimates of average or even mid-to-later-life survival.

This general lack of rank concordance does not seem to be a result of sampling noise due to smaller population sizes in extreme old age (Fig 2d-f). The growth of centenarian population sizes during the past 51 years, by several orders of magnitude, has not been marked by any appreciable increase in the rank-concordance of official mortality statistics or centenarian attainment (Fig 2e-f). In addition, the centenarian attainment of countries remains relatively stable across time: countries retain consistent rankings for late-life survival after cohorts become extinct and are completely replaced (Fig 3; Table 1; Table S1). Such long-term consistency would not occur if rankings were driven by stochastic random-sampling effects.

**Figure 3.**
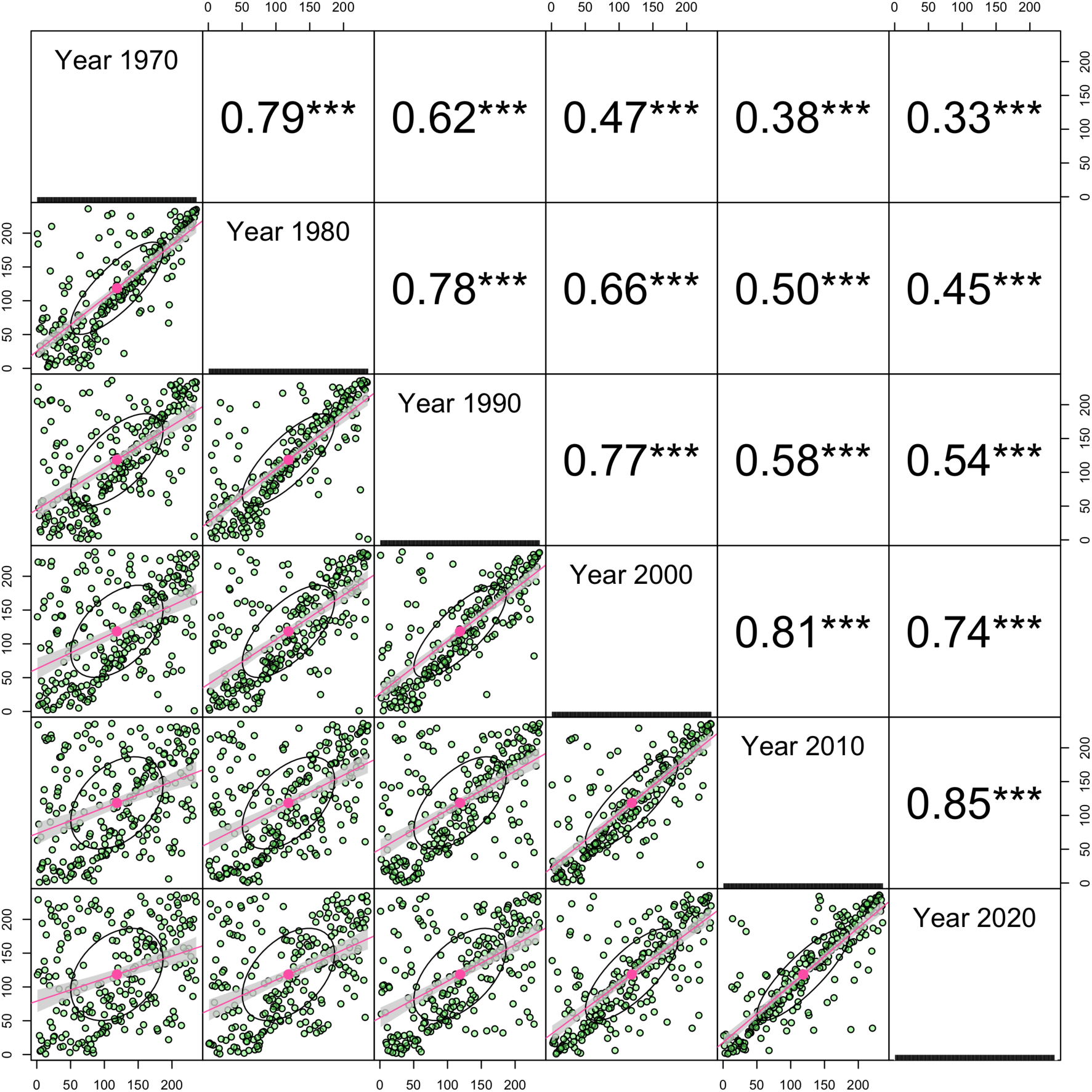
Rank concordance across non-overlapping cohorts of the oldest old. The ranks for centenarian attainment – survival rates from age 80 to ages 100+ – remain consistent across sizeable gaps and degrade slowly over time, despite the complete replacement of cohorts. Shown here are rank concordance (below the diagonal; pink lines show linear least squares regressions; ranks shown on axis labels) and correlation coefficients for rank concordance (above the diagonal) across 10-year gaps. These data all but rule out sampling effects as the dominant driver of late-life mortality rankings: virtually every 100+ year old dies and is replaced each decade, yet there is persistent long-term concordance in late-life survival over a decade or more. This concordance is typically around R^2^ = 0.8 across a one-decade gap and around half of the variance in late-life mortality remains after 30 years. This slow decay in concordance likely represents long-term shifts in underlying vital registration error rates and population health patterns.

Such patterns also did not disappear when comparing centenarian attainment rates, rather than ranks, within years (Fig S3). Beyond introducing a minor degree of zero-inflation, analysing unadjusted data did not appreciably alter the patterns seen in rankings. That is, unadjusted late-life data reinforces the observation of a persistently weak or non-existent relationship between late-life and earlier-life survival (Fig 2; Fig S3): a pattern that is sustained long-term, despite dramatically increasing sample sizes over time (Fig 2d).

Our cohort estimates of centenarian attainment and late-life survival assumed closed cohorts based on the negligible migration rates of many 70+ populations^32^. A key shortfall of estimating the centenarian attainment rate was, therefore, the lack of age-specific migration data available to test this assumption.

There are two key factors that mitigate this issue. First, it seems reasonable to assume that migration pressures would generally act to increase the advantage of states that had higher late-life (ages 80+) survival rates, as these would generally be regions with better health and social aged care. Inversely, it seems reasonable to assume that regions with high mortality rates in over-80 populations would not attract substantial net inflows of late-life migrants. Migration pressures should, therefore, act to amplify any differences in centenarian attainment between healthy and unhealthy populations, without substantially changing centenarian rankings. The second factor, that would again mitigate this issue, is the strikingly low rate of global migration in the over-80 population. Less than 7% of the over-80 population are international migrants^34^ at any point during their lives, and almost none of that migration takes place at an advanced age^32^, limiting the possible effect of net migration flows when estimating cohort mortality above age 80.

There may be partial exceptions to this rule in some regions. It is possible, for example, that the extraordinarily high estimate for centenarian attainment rate of Monaco may instead result from a net inflow of wealthy European old-age migrants^35^ who are drawn to the zero rate of Monégasque income, wealth, inheritance, property taxes, avoiding the high inheritance or death taxes in nearby home countries like France.

Although the low migration rate of over-80s makes this unlikely, similar (state-internal) migrant flows of over-80 retirees may also partially explain the higher centenarian attainment calculated for current and former colonial holdings such as Guadeloupe and New Caledonia. However, a simple alternative hypothesis may be that these regions have received far less health funding, have had much less invested care in vital registration by colonial governments, and therefore maintain far less accurate government records than other regions.

Furthermore, migration does not explain the high frequency of low life-expectancy states like Malawi and Cote d’Ivoire, where a large inflow of over-80 migrants seems somewhat less than likely, or Turkmenistan which allows no migration at all. Thus, while we cannot rule out that these collective patterns are shifted or offset by migration, it seems generally unlikely that the highly irregular distribution of survival in older individuals could be neatly explained by migrant flows, given over-80 populations almost universally remain in their home country.

A much simpler explanation may be that the combination of an unrecognised error process^23^, undetectable age-coding errors^26^, and the lack of any capacity to physically validate human ages has led to a numerical fiasco.

## Discussion

Aggregation of ages in identity documents seems, in the case of UN data at least, to generate patterns at odds with rational expectations. Yet ironically it can only be asserted that such data are absurd, rather than conclusively demonstrated, for precisely the reasons detailed above and below: measuring age-coding error rates, rather than assuming or guessing their value^22,26,36^, is not possible.

It may be that demographers will find some qualitative or simulated reason to explain away why Monaco, Thailand, Western Sahara and Malawi all possess remarkably low old-age mortality rates over time. Reversion to unmeasured and unmeasurable ‘hidden’ population heterogeneity seems a likely response^37,38^. One leading demographer has already suggested, for example, the overtly racist idea that “selection of the strongest” through “the tremendous health selection effect of slavery” and “high fertility among black people”^39^ provides a way for population heterogeneity to explain the excess number of extremely old people in Martinique and Guadeloupe^39^. This does not, of course, explain why the near-equal-sized migrant populations from Martinique and Guadeloupe that reside in mainland France do not survive to remarkable ages after they emigrate to regions with better healthcare^26^, and the simplest explanation seems that age-coding errors in the overseas territories are more common than on mainland France. However, even if such ideas continue to find traction, the root cause of the debate will remain.

Documents measuring human ages are not calibrated against any biometric or physical test. Instead, the only widely-implemented approach to detect age errors has been to cross-check paperwork, a process that has been repeatedly and incorrectly referred to as validation^40–43^. Paperwork can be, and often is, both perfectly consistent and perfectly inaccurate, in the same way that always scoring a zero on a dartboard is both perfectly consistent and perfectly inaccurate. Cross-checking ages across different documents to see if they match has therefore provided an illusion of accuracy and validation where none exists^25,26,44–46^.

This lack of physical calibration for human ages places enormous trust in a reporting system that relies on paper records. Paper records are routinely incorrect^26,45–47^, fabricated for personal gain^48^, and necessarily decades old by the time they are used to measure adult ages. Compounding this problem, most documents do not report ages independently but simply duplicate the age written on another document, allowing the consistent propagation of errors both forward and, through replacement of earlier documents and birth certificates^48^, backward through (bureaucratic) time^26^. Even more remarkably, some countries have allocated birth certificates to their entire population by simply guessing the age of their citizens^49^ or, in the case of the USA, taking self-reported ages at face value when issuing birth certificates *en masse*^12,13^.

Vital registration also remains almost universally incomplete at national and subnational levels^10–12^, and even highly literate populations routinely forget, mis-record, or misreport their age when birth documents are present^50,51^. In 1960, for example, between 34% (‘nonwhite female’[sic]) and 73% (‘white male’) of US citizens reported a different age in the census to their ‘official’ age records^52^, with over 25% of non-white respondents reporting an age that was mismatched by more than 10 years^52^. Even now, US officials are instructed not to use birth certificates as evidence for age or identity^48^ because according to the US government “a birth certificate cannot be positively linked with an individual” and “most birth certificate fraud is committed using genuine documents”^48^ that contain consistently reported ages.

Such problems are echoed across other countries. For example, after being considered some of the best-quality data in the world^36,40,53^, 82% of 100+ year olds in Japan were discovered to be dead^45^ in 2010 and at least 72% of valid Greek centenarians were discovered to be the product of pension fraud^26^ in 2012. These cases were often detected, not through inconsistencies in their (often perfectly consistent) documents, but by attempting to find the physical person holding those documents and discovering they were dead^54^. The number of cases in which living people hold consistent and inaccurate documents is therefore unknown. It is not possible, or rather has not been possible^15–18^, to accurately measure age in a developmentally mature person without relying on either paperwork or guesswork. This inability to detect inaccurate data has been dramatically illustrated by the repeated document-based validation of the world’s oldest people who, after as long as a century of intensive public and expert scrutiny^46,47^, are discovered to be fake^46,47^ or dead^26,44,45^.

Yet this situation is, for the first time, becoming unnecessary. The emergence of new, paperwork-independent metrics of human age presents a solution to measuring, rather than guessing or assuming, the error rate of document-derived age data. Algorithmic age estimates can be now derived from facial images and epigenomic diversity^15,16,18^.

When epigenomic age estimators have been applied to ‘supercentenarians’ (individuals aged 110+), centenarians (100+), or the oldest-old (80+), they have almost universally indicated that these individuals’ ages are substantially younger than their documents suggest^55–59^. This is even the case when different epigenomic clocks, driven by variation at thousands of unlinked loci, have all independently suggest that the paperwork-ages are over-estimates^55^.

This has somehow led most ageing researchers to conclude that these people must enjoy some kind of near-superhuman advantage in slowing the ageing process which, somehow, consistently alters the thousands of (presumably non-causal) variables used to construct epigenomic clocks^55–59^. While this might be possible, although statistically extraordinary, a far simpler alternative is that the age measured by epigenomic clocks is right and the paperwork is wrong. This solution, regrettably, does not yet seem to have occurred to many anti-aging enthusiasts working in the field^55–59^.

Adding a third independent measurement based on unbiased physics-based testing would resolve which of these estimates of age— the paperwork, or the epigenome – are correct. Not only are such tests possible^60,61^, they would present an exciting and simple way to resolve the deep problems of age validation in demography.

The 25% of children living without a birth certificate today^62^, and the much higher fraction of adults who lacked documents to officially validate their ages^10,11,13^, could then be assigned ages and identity documents with a measurable degree of certainty. This would both dramatically improve the conduct of public health worldwide, allow strict enforcement of laws on child trafficking, the age of consent, and the human rights of the child, and resolve the contentious nature of extreme-age records. Given the potential payoffs and low cost, some thought should be allocated to testing such a solution.

## Methods

Raw and processed population data were downloaded from the United Nations World Populations Prospects^27^ (Supplementary Code). As centenarians are typically less than 0.1% of the population^27^, factors that modify population size in the near term and which primarily affect younger populations are capable of dramatically distorting the per-capita rate of centenarians. Raw cross-sectional per-capita rates, a simple count of the number of centenarians per person, are therefore highly susceptible to distortion form population growth and migration amongst younger people and were ignored as an indicator of centenarian density.

Therefore, to avoid distortions driven by earlier-life migration and survival, cohorts were linked longitudinally from a baseline at age 80-84, until reaching the terminal 100+ age category, and mortality rates calculated across this 20-year period by dividing the resultant age 100+ population by the initial population exposed to risk at age 80-84. We termed this late-life cohort survival rate the centenarian attainment rate.

Calculations of centenarian attainment made two key assumptions: that net migration above age 85 was negligible, and that survival of individuals to age 105+ was sufficiently rare and random compared to 100-104-year-olds that their pooling into this cohort introduced a negligible degree of upward bias in apparent survival rates.

The former negligible-migration assumption is supported by UN estimates of the percentage of the over-75 population born overseas, *i.e.* who are international migrants that migrated at any time in their lives, which is below 7.2% every year^34^ from 1990 to 2020. This upper estimate is likely driven by migration at much younger ages, a likelihood reinforced by UN observations and models that suggest migration rates fall dramatically after a small post-retirement ‘bump’ in a few countries^32^ and are effectively zero beyond age seventy or eighty^32^. However, diversity between states is potentially high as noted in the results and discussion.

The latter assumption is generally untestable as estimates of extreme late-life survival are rarely available within countries, and existing estimates routinely appear to involve researcher incompetence^63–66^, deeply questionable model choices^24,67^, and even hiding inconvenient data above the y-axis^68^. Thus, while some isolated estimates place the ratio of 105+ year olds to 100+ year olds within a cohort at around 2-3%, such estimate are insufficiently reliable for modelling.

## Data availability statement

Data in this study are available from the World Populations Prospects database (2022 release) and are also provided in the Supplementary Materials and reproducible code.

## Code availability statement

All code is available in the supplementary materials, from the github repository https://github.com/SaulNewman/UNcentenarian and on request from the corresponding author.

## Data Availability

Data in this study are available from the World Populations Prospects database (2022 release) available at https://population.un.org/wpp/Download/Standard/CSV/ and are also provided in the Supplementary Materials and reproducible code.

## Acknowledgements

SJN would like to thank Dr Elena Racheva for her warm support.

## Author contributions

SJN conceived, designed, preregistered, coded, wrote, and edited the manuscript and constructed the figures.

---

**Figure S1.**
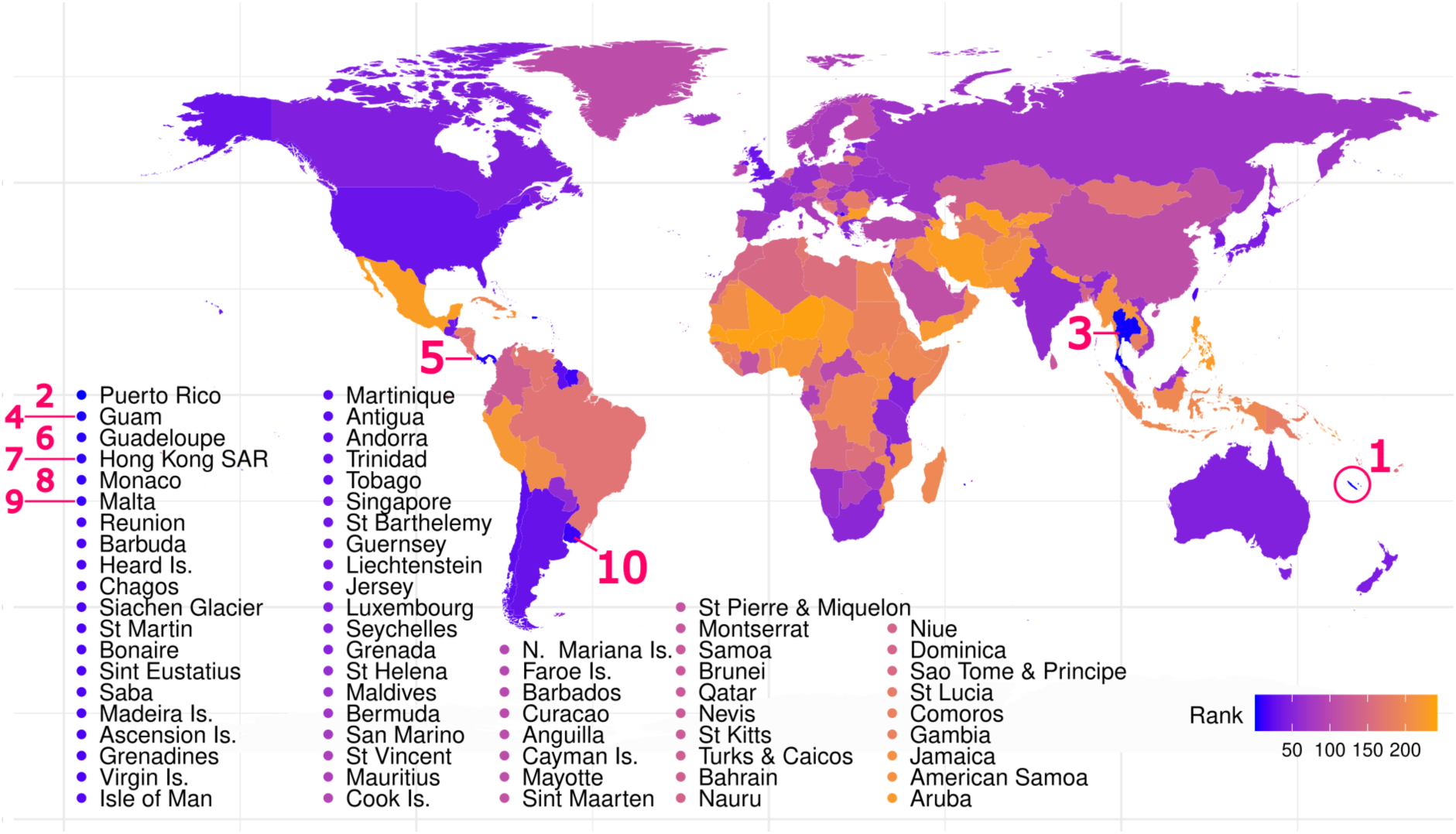
Patterns of UN-estimated life expectancy at age 100. World rankings of life expectancy at age 100, in years, derived from the 2021 UN World Population Prospects data^27^. Rankings for the ten longest-lived populations at age 100 are numbered.

**Figure S2.**
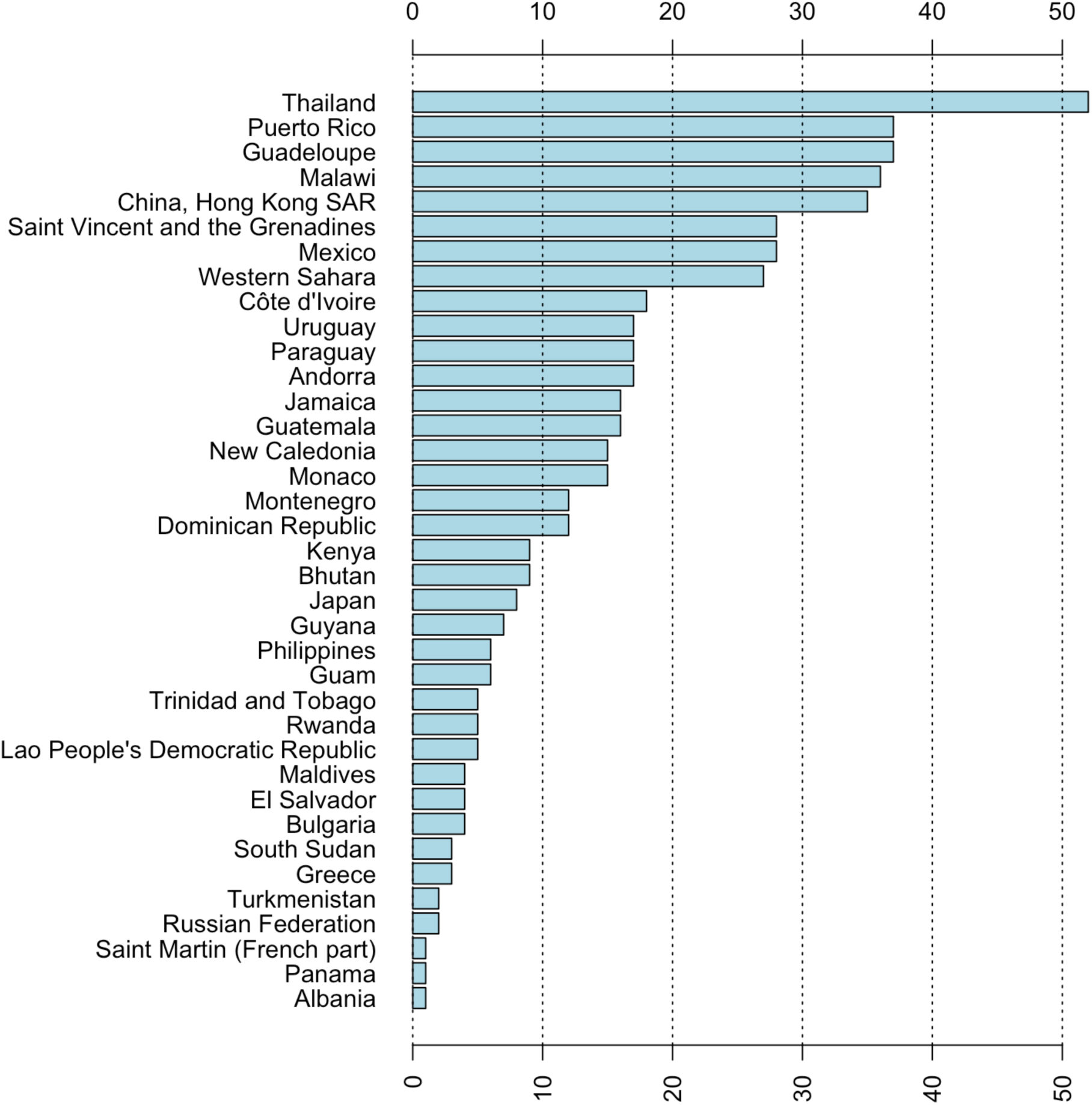
Incidence of top 10 rankings for centenarian attainment across 1970-2021. Despite the seemingly highly random composition of countries that attain the most remarkable late-life survival across the globe, rankings across consecutive cohorts are highly consistent. This is evident in the frequency with which several countries have retained top 10 rankings for late-life survival across the 51 years of available UN data. Thailand occupies a top 10 spot every year, closely followed by Puerto Rico, Guadeloupe, and Malawi with over 30 appearances each. Some countries, such as the Russian Federation, fell out of the rankings when they cease to exist, while others like South Sudan are recently created states. This ranking consistency rules out stochastic or sampling processes as a cause of these rankings, as age 100+ populations undergo complete replacement every 5-10 years.

**Figure S3.**
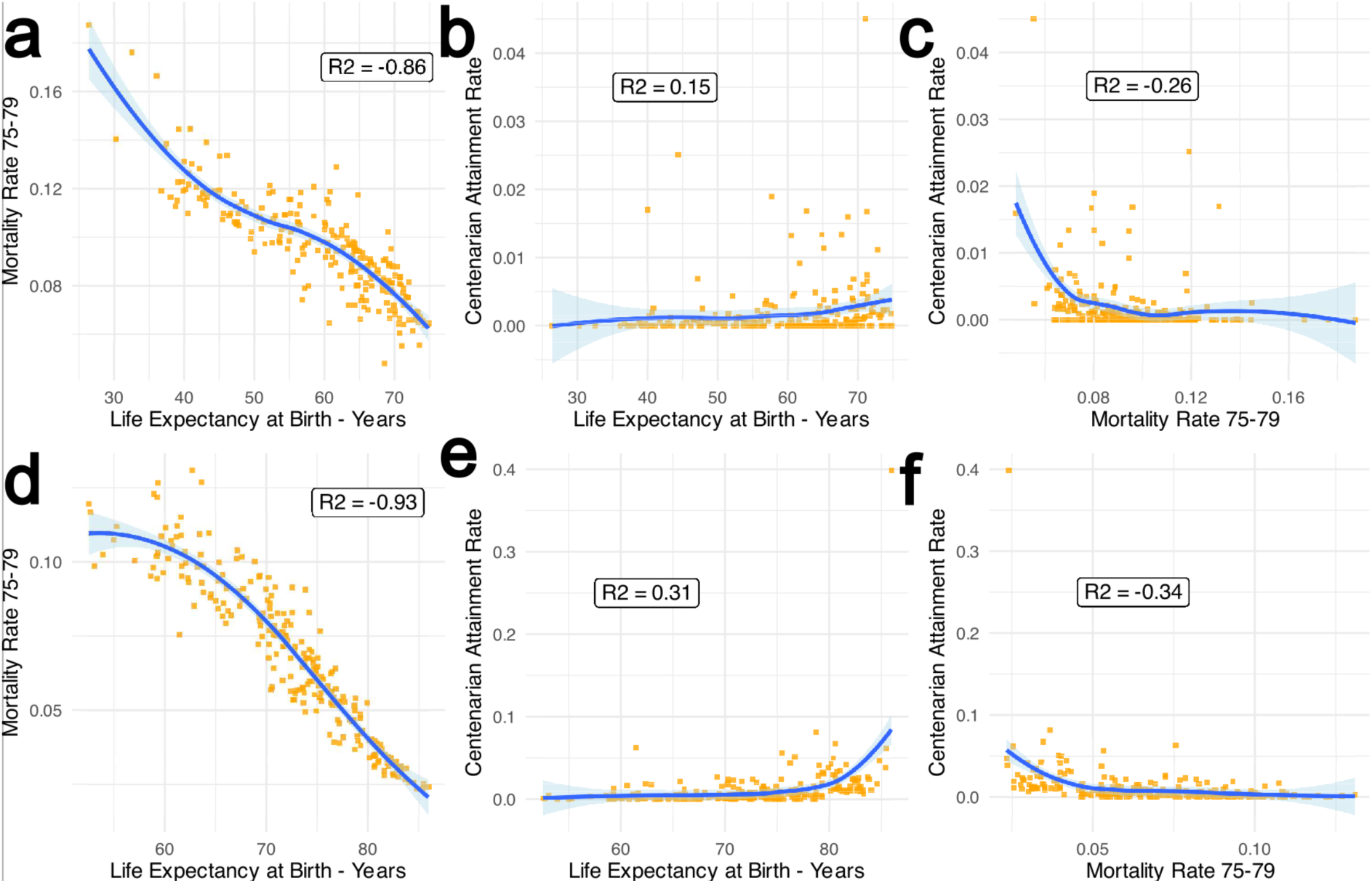
Consistently weak relationships between late-life survival or centenarian attainment rates and earlier-life survival over time. World Population Prospects data reveal the persistently weak relationship between early-life survival and centenarian attainment rates from the earliest (a-c) to the latest (2021 data, d-f) available mortality data. As expected, life expectancy at birth is strongly negatively correlated with age-specific mortality rates until age 75-79 (a,d). Life expectancy at birth calculations include of survival rates from ages 80-100+ and should, therefore, be highly negatively correlated. However, life expectancy at birth then has a persistently weak relationship with late-life survival and centenarian attainment rates from 1970 (b) to 2021 (e) despite orders-of-magnitude growth in the size of older populations. Even mortality rates at age 75-79, immediately before ages 80-100+, display persistently weak correspondence with centenarian attainment rates (c,f). Such correlations between early- and late-life survival appear to be largely driven by a few low-mortality countries at the margin of the survival distribution (Supplementary Code; blue lines show locally weighted smoothed splines).

**Table S1.** The United Nations top 10 countries for life expectancy at age 100+, in years, from 1970-2021 during select years.

| Life expectancy at age 100 in years - Country |  |  |  |  |
| --- | --- | --- | --- | --- |
| Rank | 1950 | 1960 | 1970 | 1980 |
| 1 | 3.0 - Bulgaria | 2.9 - Thailand | 3.5 - SVG | 3.7 - Thailand |
| 2 | 2.6 - Thailand | 2.8 - Puerto Rico | 3.3 - Thailand | 3.6 - SVG |
| 3 | 2.4 - Portugal | 2.8 - Bulgaria | 2.9 - Puerto Rico | 2.9 - N. Caledonia |
| 4 | 2.4 - Guatemala | 2.8 - SVG | 2.8 - Bulgaria | 2.9 - Puerto Rico |
| 5 | 2.4 - SVG | 2.6 - Guyana | 2.7 - Guyana | 2.6 - Guadeloupe |
| 6 | 2.2 - Ireland | 2.4 - Ukraine | 2.4 - N. Caledonia | 2.5 - Montenegro |
| 7 | 2.1 - Panama | 2.3 - Lithuania | 2.3 - Belize | 2.5 - Belize |
| 8 | 2.1 - USA | 2.3 - Belarus | 2.3 - Hong Kong | 2.5 - Guyana |
| 9 | 2.1 - Türkiye | 2.2 - Grenada | 2.3 - Grenada | 2.4 - Uruguay |
| 10 | 2.1 - Iceland | 2.2 - Belize | 2.2 - Montenegro | 2.4 - Panama |
| Rank | 1990 | 2000 | 2010 | 2021 |
| 1 | 4.2 - Thailand | 3.7 - Thailand | 3.8 - N. Caledonia | 4.3 – N. Caledonia |
| 2 | 3.4 - SVG | 3.2 - Uruguay | 3.7 - Thailand | 4.3 – Puerto Rico |
| 3 | 3.4 - Montenegro | 3.1 - N. Caledonia | 3.4 - Guam | 4.1 - Thailand |
| 4 | 2.8 - Puerto Rico | 3 - Puerto Rico | 3.4 - Uruguay | 3.9 - Guam |
| 5 | 2.8 - N. Caledonia | 3 - El Salvador | 3.2 - Puerto Rico | 3.2 - Panama |
| 6 | 2.7 - Uruguay | 2.9 - Guadeloupe | 3.2 - Guadeloupe | 3.2 - Guadeloupe |
| 7 | 2.7 - Guadeloupe | 2.9 - Gibraltar | 3.0 - Monaco | 3.1 – Hong Kong |
| 8 | 2.6 - Bahamas | 2.8 - Guam | 2.9 - SVG | 3.0 - Monaco |
| 9 | 2.5 - Guyana | 2.7 - Bahamas | 2.9 - Gibraltar | 3.0 - Malta |
| 10 | 2.5 - Gibraltar | 2.7 - Monaco | 2.8 - Suriname | 3.0 - Uruguay |

Abbreviations: SVG = Saint Vincent and the Grenadines; N. Caledonia = New Caledonia; Hong Kong = Hong Kong Special Administrative Region

